# Differences in Declines in Pediatric ED Utilization During the Covid19 Pandemic by Socioeconomic Disadvantage

**DOI:** 10.1101/2021.04.09.21255225

**Authors:** Bisakha Sen, Anne Brisendine, Pallavi Ghosh

## Abstract

**Background:** There is growing evidence that the early months of the COVID19 pandemic saw substantial declines in pediatric Emergency Department (ED) utilization in the U.S. However, less is known about whether utilization changed differentially for children who are socio-economically disadvantaged. We examined how changes pediatric ED visits during the early months of the COVID19 pandemic differed by two markers of socio-economic disadvantage, minoritized race and being publicly insured.

**Methods:** This retrospective observational study used electronic medical records from a large pediatric ED in a Deep South state for January-June 2020. Three time-periods ╌ pre-COVID19 (TP0), COVID19 with restrictions like stay-at-home (TP1), and COVID19 with restrictions relaxed (TP2) in 2020 were compared with the corresponding time-periods in 2019. Changes in overall visits, visits for minoritized race (MR) versus non-Hispanic white (NHW) children, and Medicaid-enrolled versus privately-insured children were considered, and changes in acuity-mix of ED visits and share of visits resulting in inpatient admits were inspected.

**Results:** Compared to 2019, total ED visits declined in TP1 and TP2 of 2020 (54.3%, 48.9%). Declines were larger for MR children (57.3%, 57.8%) compared to NHW children (50.5%, 39.3%), and Medicaid enrollees (56.5%, 52.0%) compared to the privately insured (48.3%, 39.0%). Particularly, MR children saw steeper percentage declines in high-acuity visits and visits resulting in inpatient admissions compared to NHW children. The mix of pediatric patients by race and insurance-status, as well as the share of high-acuity visits and visits with inpatient admissions differed between TP1 and TP2 of 2019 and 2020 (p<0.05 for all cases). In contrast, there was little evidence of difference between TP0 of 2019 and 2020.

**Conclusion:** The role of socioeconomic disadvantage and the potential impacts on pediatric ED visits during COVID19 in the Deep South of the United States changes is understudied. We find evidence of steeper declines in visits among MR and Medicaid-enrolled children, including for high-acuity conditions, than their NHW and privately-insured counterparts. Since disadvantaged children sometimes lack access to a usual source of care, this raises concerns about unmet health needs, and worsening health disparities, in a region that already has poor health indicators.

## Introduction

The COVID19 pandemic has brought about seismic disruptions to societies and to healthcare systems around the world. In the United States, a national emergency was declared on March 13, 2020 in response to COVID19. Subsequently, many states instated measures like closing schools and businesses and issuing ‘stay at home’ orders of varying degrees of stringency. Further, the Centers for Disease Control and Prevention (CDC) and the Centers for Medicare & Medicaid Services (CMS) recommended delaying elective care and prioritizing urgent care; nonetheless the early months of the pandemic saw sharp declines in Emergency Department (ED) visits in the U.S. [1-4] including ED visits among the pediatric population [5-7]. The decline was likely driven by fear of COVID19 infection and perhaps exacerbated by access issues due to lockdowns and stay-at-home orders [4, 8, 9].

Some of the decline in pediatric ED visits could be due to reduced needs – for example, lockdowns and school closures could have resulted in fewer vehicular injuries or fewer infections or injuries incurred at school. However, existing studies show that ED visits for serious health conditions declined for both adults [3] and for children [7], raising concerns about unmet health needs that may lead to subsequent adverse effects. Further, while the decline in pediatric ED visits has been recorded using data from single EDs [8, 10] to multiple EDs across several states [1, 5, 7], one issue that is relatively unexplored is whether declines differed for socioeconomically disadvantaged children, particularly children from communities of color that have been disproportionately impacted both in terms of illness and fatality from the virus and the economic fallout of the pandemic. This is particularly relevant since existing research shows that minoritized race and ethnicity, public insurance and low-income status are predictors of using the ED for healthcare needs in lieu of a usual source of care [11].

We conducted a retrospective, observational study to investigate the change in pediatric ED volume during the early months of the pandemic in the largest pediatric ED facility in the state of Alabama. Alabama is one of the Deep South states ╌ characterized by large African-American populations, high poverty rates, and low ranking on health indicators. Alabama and other Deep South states like Mississippi, Georgia and Alabama are among the states that had the highest number of ED visits in 2019 in the U.S. [1]. Yet, to our knowledge, most existing studies on changes in pediatric ED visits during COVID19 have not included data from the Deep South, leaving an important gap in this literature.

The primary indicator of socio-economic disadvantage that we considered was minoritized race (MR) as opposed to non-Hispanic white (NHW). We also considered Medicaid enrollment as a proxy measure of low family income. We further considered changes in visits of different acuity levels and visits that resulted in inpatient admissions, and whether the extent of these changes differed by race and by insurance status. Finally, we compared changes in the early months of the pandemic when state-imposed restrictions such as stay-at-home orders were in place, versus the subsequent months when these restrictions were largely relaxed.

## Methods

We used Electronic Medical Records (EMR) data for all pediatric patients presenting to the ED from January 1^st^ through June 15^th^ of 2020, and for the same time-period in 2019. We extracted the following information: (i) date of visit; (ii) patient’s race-ethnicity; (iii) the patient’s insurance status; (iv) acuity level of the patient and (v) whether the ED visit resulted in an inpatient admission.

For analysis purposes, our main variable of interest were recoded as follows: Race and ethnicity was categorized as the binary indicator NHW versus MR; the latter primarily consisted of African-American patients, but also included Hispanics and other races since they were too small in number to permit a separate category. Subsequently sensitivity analyses that excluded the “other race” category were conducted and found no impact on the results. Insurance status was categorized as private, Medicaid (which covered children up to 141% of the federal poverty level); ALL Kids, ╌ Alabama Children’s Health Insurance Program (which covered children from above 141% to 312% of federal poverty level); self-insured /not reported; and out-of-state. Acuity-level was originally denoted on a 5-point scale, with level 1 denoting most urgent and resource-intensive to level 5 denoting the least urgent, and two separate categories (T1 and T2) for patients presenting with trauma. It was recoded for analysis purposes into three categories: ‘high-acuity’ which included levels 1 and 2 and the two categories of trauma, ‘mid-acuity’ which included level 3, and ‘low-acuity’ which included levels 4 and 5 of the 5-point scale. Inpatient admission was coded as a binary indicator of whether the ED visit resulted in an inpatient admission or not.

We compared patients presenting at the ED in three time periods in 2020 to those presenting during the same periods in 2019. January 1-March 15 is time-period ‘0’ (TP0). In 2020 this was the time-period when COVID19 cases were essentially undetected in Alabama, and no restrictions had been recommended or required, thus the comparison of TP0 in 2019 with TP0 in 2020 serves as a ‘counterfactual’ (or hypothetical, alternative state in which there was no pandemic) because no changes due to COVID19 should be seen in this period of 2020. March 16-April 30 is time-period ‘1’ (TP1). In 2020, this was the pandemic period when restrictions were also in place in Alabama. For example, limits on businesses and large gatherings started on March 16, public spaces and beaches closed on March 18, the largest city and county started issuing stay-at-home orders on March 24, and statewide stay at home orders were imposed on April 3.[12]. May 1-June 15 is ‘time-period 2’ (TP2), which, in 2020 was the pandemic period when restrictions relaxed or lifted. Non-emergency medical procedures were permitted starting April 28, and from May 1 onwards businesses, restaurants, salons, parks, and beaches were allowed to start re-opening though a ‘safer at home’ advisory remained. The population was asked to voluntarily adopt safety measures, but ‘mask mandates’ were not introduced at a county or state level until July 2020, which is after our study period.

We presented data from the three time periods from 2019 and 2020 to illustrate changes in ED visits for the full sample, by MR status and insurance status, and for acuity and inpatient admission for MR and NHW children. Thereafter, we used chi-square analyses to test whether the distribution of these characteristics changed for pediatric patients presenting to the ED in 2020 versus the same time period in 2019. Statistical significance was set at P<0.05.

All analyses were completed using STATA version 16. The study protocol was approved by the Institutional Review Board of [BLINDED FOR REVIEW – University name and IRB Registration Number] as exempt. No informed consent was obtained for the study as the data were retrospective medical records and were de-identified prior to receipt by the authors. The dataset generated and analysed during the current study are not publicly available due to hospital and IRB policies but may be available from the corresponding author on reasonable request.

## Results

There were 56,550 total ED visits in our combined study periods, of which 26,683 were NHW and 29,867 were MR children. Of the latter group, 96.9% were African-American, 0.9% were Hispanic, and 2.2% were other races, bi-racial or unreported. Insurance type differed significantly by race (p<0.01) ╌ among MR children 76.9% were insured by Medicaid, 4.9% by ALL Kids, 13.1% were privately insured, and 3.0% were uninsured or out of state or unknown; for NHW children, corresponding figures were 49.3%, 6.6%, 41.1%, and 4.1%. Of the overall sample, 52,0% were males and 48.0% females.

Table 1 shows total visits to the ED for the three time periods in 2019 and 2020, and distributions by race, acuity level, inpatient admits and insurance-status. In TP0 2020, 15,182 pediatric patients presented to the ED compared to 15,725 in TP0 2019 – a decline of 3.4%. In contrast, in TP1 2020, 4,093 patients presented for care, a decline of 54.3% from 8,950 patients in TP1 2019; and in TP2 2020, 4,258 patients presented, a decline of 48.9% from 8,342 patients in TP2 2020. Compared to 2019, MR patient visits declined by 2,690 (57.8%) in TP1 of 2020 and 2,570 (57.3%) in TP2 of 2020. In contrast, NHW patient visits declined by 2,167 (50.5%) in TP1 and by 1,513 (39.3%) in TP2. The magnitude of declines differed across acuity-level. In 2020 as compared to 2019, low-acuity visits declined from 5,570 to 2,272 (59.2%) in TP1 and from 5,361 to 2,230 (58.4%) in TP2; high-acuity visits declined from 1,397 to 744 (46.7%) in TP1 and 1,185 to 774 (34.7%) in TP2. Visits resulting in inpatient admissions declined from 1285 to 828 (35.6%) in TP1 and from 1,159 to 906 (21.8%) in TP2. Finally, compared to 2019, the number of Medicaid-enrolled patients presenting to the ED declined by 3,204 (56.5%) in TP1 and 2,792 (52.9%) in TP2 of 2020, whereas number of privately-insured patients declined by 1,148 (48.3%) in TP1 and by 869 (39.0%) in the same periods. Unsurprisingly, while the number of out-of-state patients was small in 2019, the percentage decline for this group was especially high in TP1 and TP2 of 2020.

**Table 1.** Distributions of Sample Characteristics in 2019 and 2020.

|  | Total |  |  |  |  |  |  |  |  |  |  |  |
| --- | --- | --- | --- | --- | --- | --- | --- | --- | --- | --- | --- | --- |
|  | T0 (Jan 1-March 15) |  |  |  | T1 (March 16-April 30) |  |  |  | T2 (May 1-June 15) |  |  |  |
|  | 2019 |  | 2020 |  | 2019 |  | 2020 |  | 2019 |  | 2020 |  |
|  | N | % | N | % | N | % | N | % | N | % | N | % |
| Total | 15,725 |  | 15,182 |  | 8,950 |  | 4,093 |  | 8,342 |  | 4,258 |  |
| Race |  |  |  |  |  |  |  |  |  |  |  |  |
| Non-Hisp. white | 7,090 | (45.1) | 6,974 | (45.9) | 4,295 | (48.0) | 2,128 | (51.9) | 3,855 | (46.2) | 2,341 | (55.0) |
| Minoritized race<sup>1</sup> | 8,635 | (54.9) | 8,208 | (54.1) | 4,655 | (52.0) | 1,965 | (48.1) | 4,487 | (53.8) | 1,917 | (45.0) |
|  | p = 0.13 |  |  |  | p <0.001 |  |  |  | p <0.001 |  |  |  |
| Acuity<sup>2</sup> |  |  |  |  |  |  |  |  |  |  |  |  |
| High-acuity | 2,397 | (15.3) | 2,337 | (15.4) | 1,397 | (15.6) | 744 | (18.2) | 1,185 | (14.2) | 774 | (18.2) |
| Mid-acuity | 2,991 | (19.0) | 3,088 | (20.4) | 1,975 | (22.1) | 1,074 | (26.3) | 1,790 | (21.5) | 1,252 | (29.4) |
| Low-acuity | 10,329 | (65.7) | 9,749 | (64.3) | 5,570 | (62.3) | 2,272 | (55.6) | 5,361 | (64.3) | 2,230 | (52.4) |
|  | p <0.01 |  |  |  | p <0.001 |  |  |  | p <0.001 |  |  |  |
| Inpatient Admit |  |  |  |  |  |  |  |  |  |  |  |  |
| Admission | 2,103 | (13.4) | 2,122 | (13.4) | 1,285 | (14.4) | 828 | (20.2) | 1,159 | (13.9) | 906 | (21.3) |
| No Admission | 13,622 | (86.6) | 13,060 | (86.0) | 7,665 | (85.6) | 3,265 | (79.8) | 7,183 | (86.1) | 3,352 | (78.7) |
|  | p = 0.12 |  |  |  | p < 0.001 |  |  |  | p <0.001 |  |  |  |
| Insurance |  |  |  |  |  |  |  |  |  |  |  |  |
| Medicaid | 10,295 | (65.5) | 9,947 | (65.5) | 5,673 | (63.4) | 2,469 | (60.3) | 5,271 | (63.2) | 2,479 | (58.2) |
| ALL Kids<sup>3</sup> | 871 | (5.5) | 874 | (5.8) | 510 | (5.7) | 221 | (5.4) | 505 | (6.1) | 238 | (5.6) |
| Out of State | 47 | (0.3) | 44 | (0.3) | 37 | (0.4) | 7 | (0.2) | 37 | (0.4) | 5 | (0.1) |
| Private | 3,904 | (24.8) | 3,776 | (24.9) | 2,377 | (26.6) | 1,229 | (30.0) | 2,227 | (26.7) | 1,358 | (31.9) |
| Self-Pay/Unknown | 608 | (3.9) | 541 | (3.6) | 353 | (3.9) | 167 | (4.1) | 302 | (3.6) | 178 | (4.2) |
|  | p = 0.62 |  |  |  | p <0.001 |  |  |  | p <0.001 |  |  |  |

Note: The strictest limitations on gatherings, businesses, and travel including a statewide stay-at-home order were in place in Alabama between March 16 – April 30, 2020 (T1, 2020). T0, 2020 had no restrictions and T2, 2020 represents the “safer-at-home” period, during which restrictions were relaxed. No statewide mask mandate existed during any of these periods.
<sup>1</sup> This category includes all patients not categorized as non-Hispanic white.
<sup>2</sup> High-acuity includes those categorized as 1 or 2 on the EMR acuity score. Mid-acuity includes category 3. Low –acuity includes categories 4 and 5 on the scale.
<sup>3</sup> Alabama’s Child Health Insurance Program (CHIP)

The uneven decline by race, acuity and insurance status contributed to a change in the distribution of patient characteristics between 2019 and 2020. From TP1 2019 to 2020, the share of NHW patients increased from 48.0% to 52.0% (p<0.05); the share of privately insured patients increased from 26.6% to 30.0% while the share of Medicaid patients fell from 63.4% to 60.3% (p<0.05); the share of low-acuity visits fell from 62.3% to 55.6% and the share of visits resulting in an inpatient admit increased from 14.4% to 20.2% (p<0.05). Similarly, between TP2 2019 and 2020, the share of NHW patients increased (p<0.05), the share of privately-insured patients increased while the share of Medicaid patients declined (p<0.05), the share of low-acuity visits fell (p<0.05) while the share of visits resulting in an inpatient admit increased (p<0.05). In contrast, there were no statistically significant changes in the distribution of patient race-ethnicity, insurance status or inpatient admits in the pre-COVID19 period of TP0 of 2020 as compared to TP0 of 2019; the one exception being acuity-level where a significant difference were seen due to increases in the share of mid-acuity cases from 19.0% to 20.4% respectively.

Table 2 shows changes in ED visits overall, by acuity level and inpatient admits for NHW and MR children. Compared to 2019, NHW children presenting to the ED declined from 4,295 to 2,128 (50.5%) in TP1 of 2020, and from 3,855 to 2,341 (39.2%) in TP2 of 2020. High-acuity visits declined by 48.0% in TP1, but by just 29.7% in TP2; and visits resulting in inpatient admits declined by 34.5% in TP1 and by just 15.8% in TP2. For MR children, the overall number presenting to the ED declined by 57.8% in TP1 and 57.2% in TP2; high-acuity visits declined by 45.0% in TP1 and 40.3% in TP2, and visits resulting in inpatient admits declined by 37.4% in TP1 and 30.8% in TP2. For both groups, the share of high-acuity cases and the share of cases resulting in inpatient admissions were higher in TP1 and TP2 of 2020 compared to 2019. There were no statistically significant differences in these shares between TP0 2019 and TP0 2020 except for the distribution of acuity level for MR children where mid-acuity visits increased from 11.8% to 13.1%.

**Table 2.**
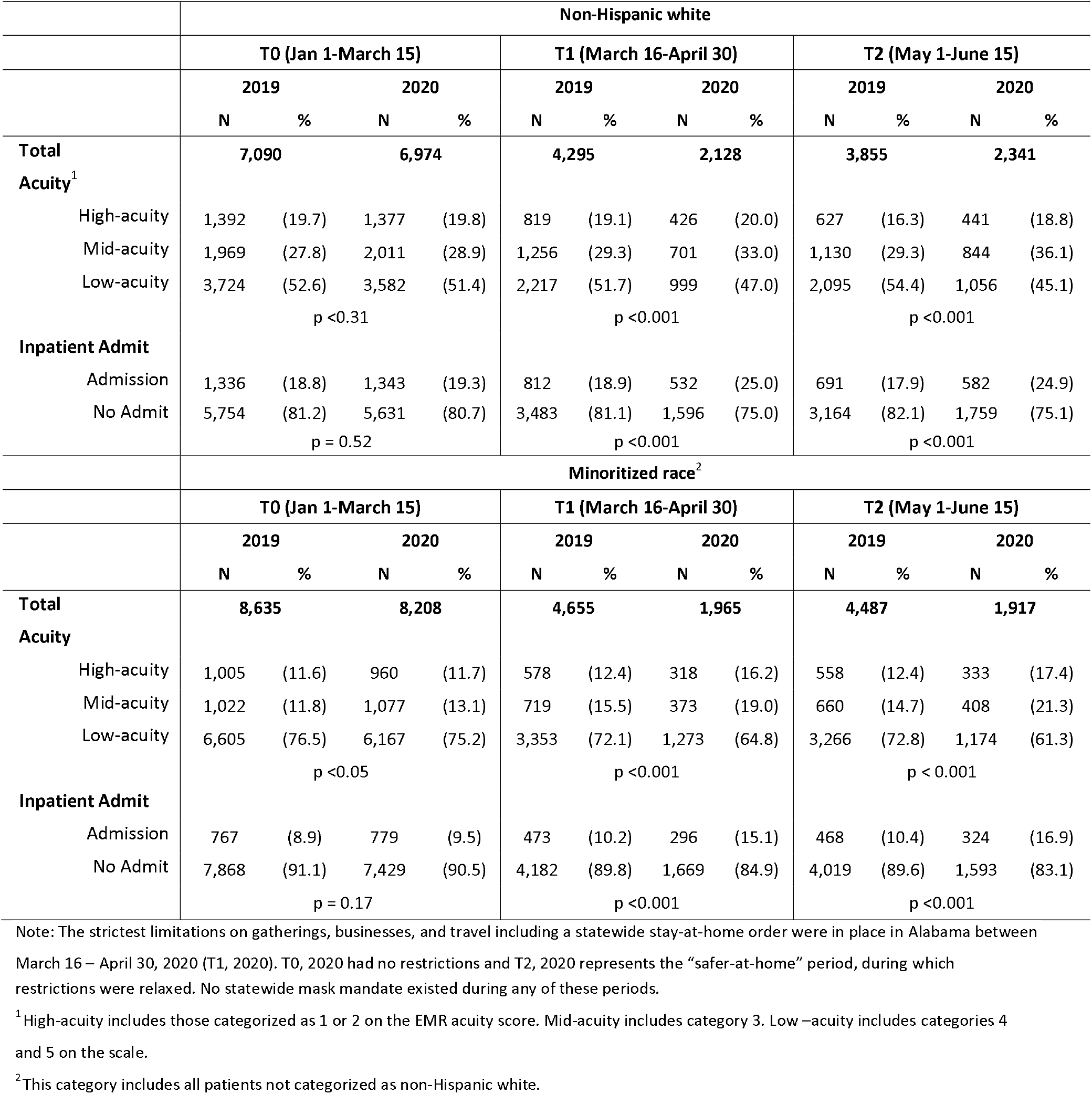
Distributions of Sample Characteristics in 2019 and 2020 by Race-ethnicity.

## Discussion

A growing body of literature has documented substantial declines in pediatric ED visits during the COVID19 pandemic in several countries [13-15]. In the US, such declines have been confirmed with data from single EDs as well as multiple EDs from several states, and overall declines as well as declines for specific health consitions have been documented [1, 5, 7, 8].

We added to this literature in several ways. First, we considered changes in pediatric ED visits by socio-economic disadvantage. The primary measure of socio-economic disadvantage was minoritized race and ethnicity, which can serve as a proxy measure for systemic and policy-based impacts on access to care and other social determinants of health. A second indicator was public insurance coverage. Communities of color and low-income communities have disproportionately borne the adverse health effects and economic effects of the pandemic [16, 17], and extant literature shows that children from these communities tend to be high users of ED for healthcare needs [11], hence exploring how their ED use was impacted in the pandemic has strong public health relevance. Second, we considered ED visits when state-level restrictions were in place versus when they were lifted. It has been speculated that restrictions like stay-at-home exacerbated the decline in pediatric ED visits, and in the Netherlands, ED visits declined by 18% in the pre-restriction period of the pandemic and by 29% after restrictions were imposed [13]. In the U.S., to our knowledge one study from Pennsylvania considered the impact of stay-at-home orders but did not include a comparison period during the pandemic when stay-at-home orders were lifted [8]. Another multi-state study indicated a ‘gradual rebound’ in visits from April but did not comment on whether this was correlated to any changes in state restrictions [7]. To be clear, we did not establish ‘causal’ links between the restrictions and visits per se – and it could be speculated that perhaps these results happened because apprehension about the virus changed over time, with NHW and relatively affluent privately-insured communities becoming less concerned. Third, we considered data from an ED in a Deep South state. The Deep South is characterized by high poverty, high shares of African-American population, lower public health rankings and higher rates of ED usage compared to the rest of US, yet few studies have considered the impact of the pandemic on pediatric ED visits in this vulnerable region.

Consistent with other studies we found that ED visits declined substantially in TP1 and TP2 of 2020 compared to 2019 – respectively 54.3% and 48.9%. However, we found substantially larger declines for MR children compared to NHW children. Furthermore, MR children did not see a rebound after state restrictions were lifted, whereas NHW children did see a partial rebound. The number of MR children presenting to the ED declined by approximately 57-58% in TP1 as well as TP2 of 2020 compared to 2019, whereas the number of NHW children declined by 50.5% in TP1, but only by 39.3% in TP2. As a result, whereas MR patients consisted of 52-55% of all ED patients in 2019 and TP0 2020, their share fell well below 50% of all patients in the pandemic period. Similar patterns are seen for publicly insured versus privately insured patients. Visits by Medicaid-enrolled patients declined by 56.5% in TP1 and 52.9% in TP2 in 2020 compared to 2019, while privately insured patients saw a decline of 48.3% in TP1 and a then a partial rebound in TP2 when the decline was just 39.0%. Also, while MR children were overall more likely to have lower acuity visits and visits that did not result in inpatient admissions than NHW children, we found evidence suggesting that MR children avoided ED visits even for more serious health conditions during the pandemic, and this continued after state restrictions were lifted. For NHW children, high-acuity visits declined by 48.0% in TP1 of 2020 compared to TP1 2019, but partially rebounded so that the decline was just 29.7% in TP2, whereas for MR children high-visits declined by 45% in TP1 and 40.3% in TP2. Similarly, visits resulting in inpatient admissions declined by 34.5% in TP1 and by just 15.8% in TP2 for NHW children but declined by 37.4% in TP1 and 30.8% in TP2 for MR children. The distributional mix of acuity and inpatient admissions was statistically different in TP1 and TP2 of 2020 compared to 2019, whereas there was no statistical difference in TP0 of 2020 versus 2019.

Prior combined data from multi-state pediatric EDs have found a decline of approximately 22% in visits for ‘serious conditions’ and a decline of approximately 43% in visits resulting in inpatient admissions. While there is a paucity of studies that stratify pediatric ED visit declines by race-ethnicity, findings from one Connecticut ED found Black children bore a disproportionate share of the decline in ED visits for mental health diagnoses as compared to white children [10]. We note that our findings regarding the racial mix of ED patients in the post-pandemic period are different from a recent study that pooled data from 27 EDs from unspecified states, and reported that the shares of NH-White, African American, Hispanic and ‘other’ patients were respectively 35.9%, 21.4%, 30.0% and 12.8% in the pandemic period, compared to respectively 32.3%, 22.3%, 33.4% and 12.0% in pre-pandemic times [5]. In contrast, we found the share of NHW patients increased to 52-55% in the pandemic period compared 46-48% in the pre-pandemic period. This further emphasizes the importance of looking at this trend of decline in pediatric ED visits by region with attention to socio-economically disadvantaged states, since otherwise critical differences may be obscured when looking at aggregated data.

Our results suggest that trends in pediatric ED visits is yet another way the COVID19 pandemic may be exacerbating health disparities. It is well documented that communities of color and low-income communities have disproportionately borne the burden of detected cases, hospitalizations, mortality and economic pain during the pandemic. Though children overall have not been as adversely impacted as older adults, there is evidence that children of color and low-income children are at higher risk of testing positive for the virus [16]. The relatively larger and persistent declines in ED visits among MR and Medicaid-enrolled children, including relatively larger declines for high-acuity visits, is likely to be directly linked to the greater disruption and greater apprehension of disease suffered by these communities. While MR and low-income are predictors of high ED usage in ‘normal’ times, often for low-acuity conditions, this is frequently because such families lack access to a usual source of care due to issues of distance, appointment availability, inability to miss work during regular hours for a medical visit, or availability of physicians willing to accept publicly insured patients. Further, ED use is often initiated because parents in disadvantaged families may have lower health literacy and are therefore less able to discern the seriousness of their child’s health conditions and are seeking reassurance [11]. Finally, EDs play a crucial role in diagnosing and treating conditions that have the potential to become severe, potentially disabling or life-threatening. There are indications that delaying ED care may have increased deaths among US adults [3, 4, 17], and that delaying pediatric resulted in ICU admissions and fatalities in Italy [6]. Further, the findings are also consistent with other indicators of delay in pediatric healthcare like vaccinations, particularly for publicly insured children.[18, 19].

We acknowledge several limitations. First, we derived our information from patient EMR data provided by the ED, hence we did not have information on reasons why patients decided to not come to the ED, whether urgent care was delayed, and whether there were subsequent adverse health consequences of the delay. Nor did we know whether and to what extent patients were able to access alternate sources of care – such as in-person or telehealth visits with healthcare providers for low acuity conditions, or direct inpatient admission arranged by healthcare providers for higher acuity conditions. Second, the Hispanic population in the state and in our sample was small, which precluded us from examining Hispanic patients as a separate category in our analyses. Third, we did not go beyond acuity-level and categorize exactly what health conditions patients were presenting with, since that has been exhaustively categorized in the multi-state studies. Fourth, while we separated our time period into the time when various state-level restrictions were in place versus not, we cannot ascribe causal impacts of the restrictions – for example, it could be speculated that the partial rebound that we saw among NHW patients was because NHW populations became less apprehensive about the virus within a few months than MR populations. Finally, while our study makes the important contribution of presenting information from a Deep South state ED, results may not be generalizable to the entire country.

In conclusion, our findings indicated that the substantial decline seen pediatric ED visits in early months of the COVID19 pandemic in the U.S. were disproportionately driven declines among racial MR and publicly insured children. Nor did these children did not see the ‘partial rebound’ in visits that their NHW and privately insured counterparts did in the latter part of this time period, that corresponded with state restrictions being relaxed. MR children also saw relatively large and persistent declines in high-acuity visits as well as visits resulting in inpatient admissions. Since MR and low-income children often lack access to a usual source of care, and ED visits may be an essential component of timely detection of health problems, this raises serious concerns about subsequent poor health outcomes and exacerbation of health disparities.

Since it is uncertain when the pandemic will abate, even after rollout of vaccines, we emphasize the need for clear communication from public health officials that is targeted to disadvantaged populations to assure the public that EDs have in place protocols to ensure the safety of their patients, and that emergency visits should not be delayed. We also emphasize the importance of continued research to understand reasons for avoiding or delaying pediatric healthcare utilization, and subsequent health consequences of that delay.

## Data Availability

The patient data used in the paper includes PHI. We are happy to share our statistical program files and detailed descriptive statistics. However, accessing the raw data will require permission from appropriate institutions.

